# Transcatheter versus Surgical Interventions for Severe Aortic Stenosis: A Contemporary Evaluation against Conservative Management

**DOI:** 10.1101/2024.04.14.24305770

**Authors:** Zhiyuan Ma, Shahrad Shadman, Yaniv Maddahi, Mahesh Krishnamurthy, Peter Puleo, Jamshid Shirani

**Author notes:** Address for correspondence: Zhiyuan Ma or Jamshid Shirani, St. Luke’s University Health Network, 801 Ostrum Street, Bethlehem, PA 18015.; (ZM); (JS).

## Abstract

**Background:** Aortic valve replacement (AVR), through transcatheter (TAVR) or surgical (SAVR) means, serves as a pivotal therapeutic approach for severe aortic stenosis (AS). While both modalities show advantages over conservative management, the long-term mortality benefits post-AVR, especially when comparing TAVR with SAVR, remain uncertain.

**Objectives:** This study aimed to perform an in-depth meta-analysis of randomized controlled trials (RCT) comparing TAVR versus SAVR, as well as their outcomes against conservative management.

**Methods:** Electronic databases were searched up to December 7, 2023. Individual patient data extracted from Kaplan-Meier plots, underwent pooling and modeling with stratification by surgical risk. The primary endpoint was all-cause mortality at 5 years.

**Results:** The study included eleven RCTs and twelve non-RCTs, encompassing 4215 patients undergoing TAVR, 4017 undergoing SAVR and comparing 11,285 AVR patients with 23,358 receiving conservative management. TAVR exhibited significantly lower all-cause mortality at 6 months (HR 0.62, 95% CI: 0.52-0.74) compared to SAVR, with no significant difference beyond 6 months (HR 1.08, 95% CI: 0.98-1.19). Additionally, over a 5-year period, there were no significant disparities in cardiovascular mortality (HR 0.98, 95% CI: 0.83-1.16) or stroke (HR 1.02, 95% CI: 0.75-1.38) between TAVR and SAVR, while TAVR exhibited a notable advantage with a markedly reduced risk of cardiovascular mortality in the initial 6 months (HR 0.64, 95% CI: 0.46-0.87) and stroke within the first month post-procedure (HR 0.31, 95% CI: 0.19-0.51). Furthermore, the mean aortic valve area and pressure gradient remained comparable between TAVR and SAVR, exhibiting stability throughout the 5-year follow-up period. AVR markedly reduced all-cause mortality compared to medical therapy (P < 0.001), with 5-year crude mortality rates of 31.6% versus 49.3%, and a difference in restricted mean survival time of 8.9 months. Similar outcomes were observed across high, intermediate, and low surgical risk categories.

**Conclusions:** While TAVR demonstrated early mortality reduction compared to SAVR, no distinctions emerged in the overall 5-year follow-up, regardless of surgical risk. AVR notably improved survival over conservative therapy. This study advocates for the preference of TAVR or SAVR in severe AS patients when feasible.

## Introduction

The rising prevalence of aortic stenosis (AS) represents a significant and escalating clinical challenge, affecting approximately 1.7% of the population aged 65 and over, with the incidence surging to 12% among those 75 years and older ^1,2^, of which 3.4% are classified with severe AS ^2^. With an aging population, the burden of AS is expected to increase further ^3^. The management of symptomatic patients with severe AS has undergone significant evolution in recent years. Surgical aortic valve replacement (SAVR) has long been the cornerstone for treatment of symptomatic severe AS, with recent guidelines expanding the indication for transcatheter aortic valve replacement (TAVR) to patients with symptomatic severe AS ranging from those at prohibitive or high risk to those at intermediate or even low risk for SAVR informed by a series of clinical trials comparing TAVR and SAVR ^4–10^. However, these trials have encountered challenges in assessing long-term mortality due to limitations in power, with many relying on composite outcomes and some prioritizing short-term all-cause mortality as the primary endpoint. Consequently, uncertainties persist regarding long-term mortality benefit comparison between patients undergoing TAVR and SAVR.

The efficacy of aortic valve replacement (AVR), whether through surgical or transcatheter approach, in treating severe AS compared to conservative management has been documented in observational studies. Several small-scale clinical trials have demonstrated the superior outcomes of TAVR over standard treatment, particularly in patients deemed prohibitive risk for SAVR ^4^ and a significantly lower incidence of all-cause death for early SAVR in asymptomatic patients with severe AS ^11,12^. Recognizing the magnitude of this benefit is crucial for decision-making in managing patients with severe AS. Despite these insights, a knowledge gap persists regarding the clinical benefit of AVR compared to conservative management in real-world clinical settings with different patient risk profiles.

In this study, we aimed to synthesize data from randomized controlled trials (RCTs) and non-RCTs to compare the outcomes of TAVR versus SAVR, as well as conservative management in patients with severe AS by pooling Kaplan-Meier–derived individual patient data (IPD) to standardize the inclusion criteria, scrutinize modelling assumption, and directly model individual-level interactions within studies, thereby enhancing statistical power and mitigating confounding bias. We sought to bridge the knowledge gap by evaluating mortality benefits in severe AS patients across various risk profiles and management strategies.

## Methods

This meta-analysis adhered to the principles delineated by the Preferred Reporting Items for Systematic Reviews and Meta-Analyses (PRISMA). The study was registered on the PROSPERO international prospective register of systematic reviews (CRD42024508950). Given its nature as a systematic review and meta-analysis, the study was considered exempt from local institutional research board review.

### Strategy of literature search for meta-analysis

We performed a systemic literature search for studies published in English after year 2000, utilizing PubMed, Google Scholar, and Cochrane Library up to December 7, 2023. The search employed keywords pertaining to severe AS and AVR (Supplementary table 1). Additionally, we also screened reference lists of eligible original studies, systematic reviews, and meta-analysis to identify any other potentially eligible studies. Subsequently, full texts of the identified studies were retrieved and thoroughly reviewed. Studies published prior to the year 2000, as well as those categorized as reviews or meta-analyses, were excluded from consideration.

### Study selection

In the analysis comparing outcomes between TAVR and SAVR, we included studies in the meta-analysis that satisfied the following criteria: 1) RCTs; 2) propensity score-matched cohort studies derived from RCTs; 3) inclusion of graphed Kaplan–Meier curves depicting clinical outcomes in the text or appendix and 4) a minimum of 1-year follow-up for outcomes. For the comparison of outcomes between AVR and conservative management, we incorporated studies that met the following criteria: 1) RCTs specifically for TAVR and SAVR with reported all-cause mortality; 2) cohort studies comparing TAVR or SAVR to conservative management; 3) cohort studies presenting reported all-cause mortality for conservative management in severe AS; 4) inclusion of graphed Kaplan–Meier curves of all-cause mortality in the text or appendix and 5) a minimum of 1-year follow-up.

### Data extraction and risk of bias assessment for meta-analysis

Two independent reviewers systematically extracted the relevant data from each study, encompassing first author, year of publication, study population characteristics (including left ventricular ejection fraction, mean aortic valve gradient and aortic valve area), study size, study design and country. Discrepancies were resolved by discussion. Raw data coordinates, comprising time, survival probability, or cumulative risks, along with the numbers at risk at specific time points and the total number of patients in each arm, were also extracted from the published Kaplan– Meier plots. The risk of bias in the included studies was assessed using version 2 of the Cochrane risk-of-bias tool for randomized trials (RoB 2) ^13^ or the Newcastle-Ottawa Scale (NOS) for cohort studies ^14^.

### Outcomes

The primary outcome of the meta-analysis was all-cause mortality at 5 years. Secondary endpoints included the composite of death from any cause, stroke or myocardial infarction (MI). Additional secondary outcomes encompassed cardiovascular mortality, and stroke or disabling stroke. The analysis also included an assessment of death from any cause specifically in patients with paravalvular leak following TAVR. The designated follow-up duration for all incorporated studies was truncated at 5 years.

### Data syntheses and statistical analysis

We reconstructed individual time-to-event data by pooling the extracted IPD using the two-stage modified iterative Kaplan–Meier approach ^15^. We used the Kaplan-Meier Method to estimate the cumulative event rate for both primary and secondary endpoints, with analysis conducted through log-rank tests. Mixed effects Cox proportional hazards models were used to assess the mortality benefit with hazard ratios (HR) and 95% confidence intervals (CI). Models were fitted with a frailty term for study-level heterogeneity. The proportional hazards assumption of Cox models for each endpoint underwent verification using Schoenfeld residuals. In case where the proportional hazards assumption was violated, two approaches were adopted. Firstly, we used landmark analysis ^16^ in which a landmark time was identified by visual inspection of the Schoenfeld residuals and Kaplan–Meier plots. The proportional hazards assumption was tested before and after the landmark time, and HRs and 95% CIs were calculated separately. Secondly, flexible hazard-based regression models were employed, utilizing by a restricted cubic spline with internal knots located at 6 and 12 months for TAVR versus SAVR, and at 12, 24 and 36 months for AVR versus conservative management. These models incorporated a time-varying treatment effect by involving an interaction term between treatment effect and baseline hazard ^17^. Trials or cohort studies were stratified into three risk groups, based on Society of Thoracic Surgeons Predicted Risk of Mortality (STS PROM) (low risk, < 4%; intermediate risk, 4-8%; and high risk, > 8%), for subgroup analysis. Studies lacking reported STS PROM were categorized based on cumulative event curves compared with other known studies. In meta-analysis of mean values, the pooling used the inverse-variance method. For studies reporting median and interquartile range, mean and standard deviation were estimated based on the sample size, median, and first and third quartiles, as previously reported ^18^. Sensitivity analyses were conducted, excluding non-clinical trials for TAVR versus SAVR or omitting studies without reported STS PROM. Additionally, the largest sample size study in the conservative management group was excluded for the AVR versus conservative management comparison. To assess the clinical advantages of AVR in comparison to conservative management, two key parameters were computed: the number needed to treat (NNT) and the restricted mean survival time (RMST). Results were presented as mean and 95% CI or ratio. The risk of outcome was quantified as HR and 95% CI. Statistical significance was determined at P values < 0.05. All analyses were conducted with R (version 4.1.2).

## Results

### Study selection and characteristics

Following a careful review of studies based on predefined inclusion and exclusion criteria, a total of 33 clinical trials and retrospective cohort studies were identified for further data extraction and analysis. This selection included multiple publications from various follow-up intervals for individual trials ^4–12,19–42^. The flowchart for the study selection is shown in Supplementary figure 1. The characteristics and quality assessment of the included studies are summarized in supplementary table 2. Of these 33 studies, there were 8 RCTs (CoreValve, NOTION, Evolut, PARTNER 1, PARTNER 2, PARTNER 3, SURTAVI, and UK TAVI) comparing TAVR with SAVR and 3 RCTs (AVATAR, PARTNER B, and RECOVERY) comparing AVR with conservative management. The risk of bias for clinical trials were assessed in Supplementary Figure 2. Baseline characteristics pooled from studies for the primary endpoint are detailed in Table 1. For TAVR (n=4215) and SAVR (n=4017), the mean age of patients was 79.5 years, with 41.1% and 44.0% being female, respectively. The mean STS-PROM scores for TAVR and SAVR were 4.8% and 4.9%, respectively. In the AVR and conservative management groups, the mean age was approximately 78.0 and 76.7, with 44.2% and 49.1% being female, respectively. The mean aortic valve area ranged from 0.72 to 0.77 cm^2^ and mean aortic valve gradient was between 45.5 and 48.5 mmHg.

**Table 1.** Baseline characteristics for trials and studies comparing all-cause mortality.

|  | Aortic valve interventions |  | Interventions versus Conservative therapy |  |
| --- | --- | --- | --- | --- |
|  | TAVR<br>(n=4215) | SAVR<br>(n=4017) | AVR<br>(n=11285) | Med Rx<br>(n=23358) |
| Age, mean (95% CI) *, year | 79.5 (76.3-82.8) | 79.5 (76.2-82.9) | (n=9432)<br>78.0 (75.6-80.5) | (n=23084),<br>76.7 (73.2-80.2) |
| Female (%) | 1732/4215 (41.1) | 1767/4017 (44.0) | 4172/9432 (44.2) | 11330/23084 (49.1) |
| NYHA III or IV (%) | 2172/3756 (57.8) | 2051/3561 (57.6) | 4427/7547 (58.7) | 224/393 (57.0) |
| Creatinine>2.0 mg/dL (%) | 69/2974 (2.3) | 59/2779 (2.1) | 186/6542 (2.8) | 70/798 (8.8) |
| Diabetes (%) | 1225/3867 (31.7) | 1196/3663 (32.7) | 2606/8510 (30.6) | 3416/14156 (24.1) |
| Hypertension (%) | 1781/2109 (84.4) | 1729/2053 (84.2) | 4130/5142 (80.3) | 8593/14156 (60.7) |
| Peripheral vascular Disease (%) | 902/3748 (24.1) | 859/3560 (24.1) | 1896/8170 (23.2) | 128/1050 (12.2) |
| Prior stroke (%) | 116/1489 (7.8) | 118/1400 (8.4) | 294/3609 (8.1) | 1176/13465 (8.7) |
| Cerebrovascular disease (%) | 512/3612 (14.2) | 485/3427 (14.2) | 1056/7259 (14.5) | 59/229 (25.8) |
| COPD (%) | 1004/3724 (27.0) | 966/3527 (27.4) | 2111/7471 (28.3) | 1626/14674 (11.1) |
| Home Oxygen (%) | 127/1517 (8.4) | 98/1480 (6.6) | 275/3217 (8.5) | 240/12358 (1.9) |
| Coronary artery disease (%) | 1643/2976 (55.2) | 1585/2827 (56.1) | 3574/6194 (57.7) | 4848/13678 (35.4) |
| Prior CABG (%) | 654/4070 (16.1) | 635/3882 (16.4) | 1393/8741 (15.9) | 192/12927 (1.5) |
| Prior PCI (%) | 713/3371 (21.2) | 662/3211 (20.6) | 1530/7522 (20.3) | 521/13078 (4.0) |
| Prior myocardial infarction (%) | 379/3472 (10.9) | 342/3305 (10.3) | 788/7566 (10.4) | 1160/13107 (8.9) |
| Pre-existing<br>pacemaker/defibrillator (%) | 406/4215 (9.6) | 395/4016 (9.8) | 849/8551 (9.9) | 51/343 (14.9) |
| A Fib/A Flutter (%) | 1099/4211 (26.1) | 1044/4012 (26.0) | 2293/9345 (24.5) | 3176/14036 (22.6) |
| STS PROM, mean (95% CI), % | 4.8 (2.0-7.6) | 4.9 (2.1-7.7) | (n=9332),<br>4.8 (3.3-6.3) | (n=1629),<br>4.4 (1.4-7.5) |
| LVEF, mean (95% CI), % | (n=2241),<br>58.7 (55.2-62.2) | (n=1992),<br>58.4 (54.3-62.6) | (n=5292),<br>60.6 (57.9-63.2) | (n=20770),<br>61.3 (57.9-64.7) |
| Aortic valve pressure gradient,<br>mean (95% CI), mmHg | (n=3531),<br>45.7 (43.2-48.2) | (n=3356),<br>45.5 (43.3-47.6) | (n=8046),<br>48.5 (45.8-51.2) | (n=20821),<br>45.5 (39.4-51.5) |
| Aortic valve area, mean (95%<br>CI), cm<sup>2</sup> | (n=3419),<br>0.75 (0.68-0.81) | (n=3221),<br>0.77 (0.68-0.85) | (n=7639),<br>0.72 (0.68-0.76) | (n=20307),<br>0.74 (0.68-0.79) |

A Fib, atrial fibrillation; CABG, coronary artery bypass graft; CI, confidence interval; COPD, chronic obstructive pulmonary disease; Med Rx, medical conservative therapy; NYHA, New York Heart Association; PCI, Percutaneous coronary intervention; SAVR, surgical aortic valve replacement; STS PROM, Society of Thoracic Surgeons Predicted Risk of Mortality; TAVR, transcatheter aortic valve replacement.
\*Mean (95% CI) was derived from meta-analysis of mean ± SD from individual study.

### All-cause mortality in TAVR versus SAVR

In this study we analyzed seven RCTs (CoreValve, NOTION, Evolut, PARTNER 1, PARTNER 3, SURTAVI, and UK TAVI) and 1 cohort study (SAPIEN 3) to compare all-cause mortality in TAVR versus SAVR. As depicted in Figure 1A, the incidence rates of all-cause mortality were comparable between TAVR and SAVR (P =0.262), with 5-year crude mortality rates of 34.0% and 34.2% respectively. Proportional hazards assumption violation was observed over the entire 60-month follow-up (P < 0.001). Time-varying HRs using flexible hazard models with restricted cubic spline indicated a relatively low HR for TAVR in the initial months compared to SAVR (Figure 1C). Landmark analyses with a 6-month cutoff maintained proportional hazards assumptions. In the initial 6 months, TAVR demonstrated significantly lower all-cause mortality with an HR of 0.62 (95% CI: 0.52-0.74) compared to SAVR. In contrast, there was no difference in all-cause mortality beyond 6 months (HR 1.08, 95% CI: 0.98-1.19) (Figure 1B and 1D).

**Figure 1.**
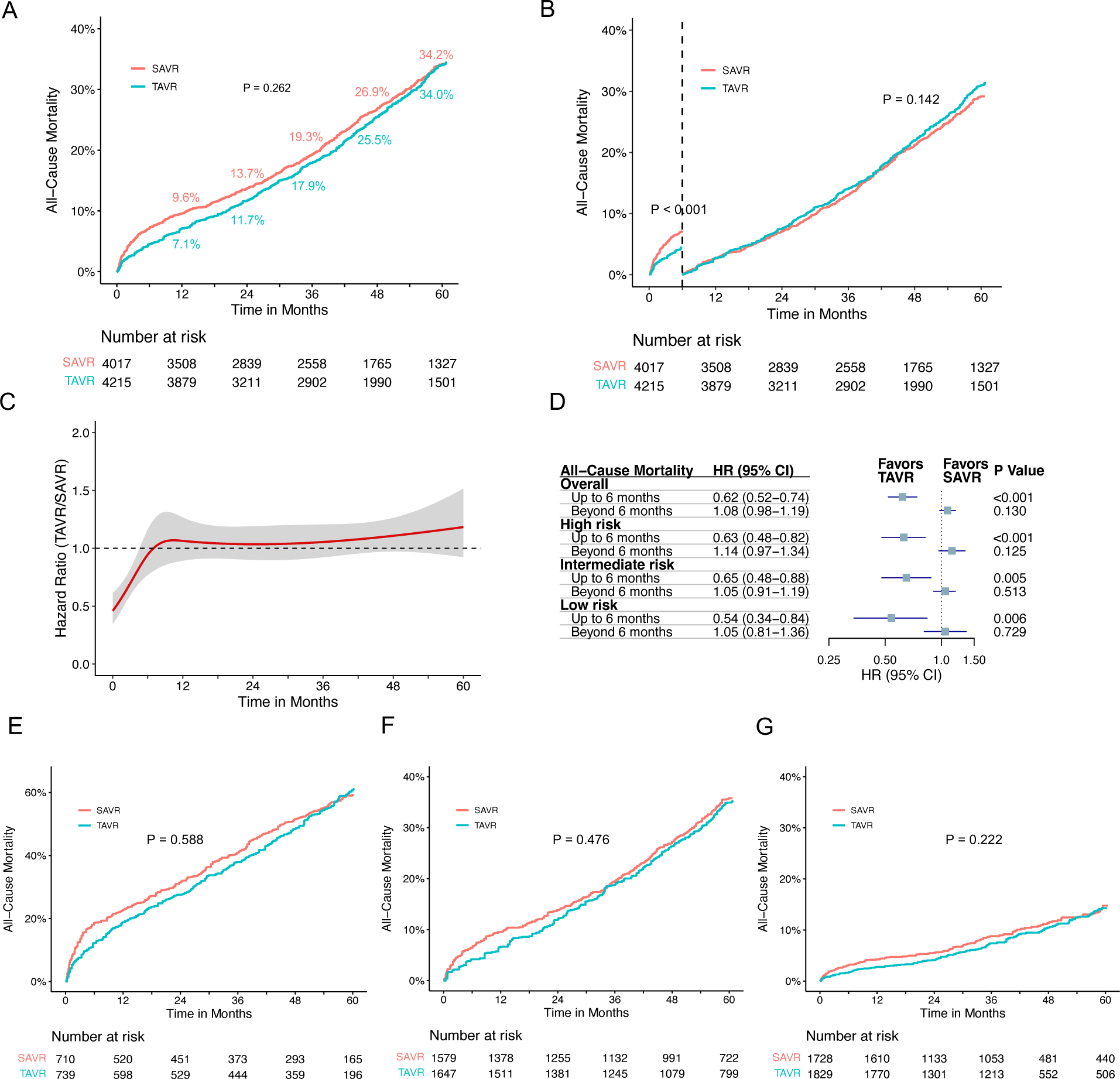
Time-to-Event Curves for all-cause mortality comparison between TAVR and SAVR. (A) Pooled Kaplan–Meier plot illustrating reconstructed IPD analysis for all-cause mortality after transcatheter and surgical AVR. (B) Landmark analysis depicting all-cause mortality for TAVR versus SAVR. (C) Hazard ratios with 95% confidence intervals depicting the dynamic change in all-cause mortality over time for TAVR compared to SAVR. The red curved line represents the time-varying hazard ratios, and the grey area denotes the 95% confidence interval. (D) Hazard ratios (HR) and corresponding 95% confidence intervals (CI) are presented for landmark analysis, both for the overall patient cohort and subgroups categorized by different risk profiles. (E) Pooled Kaplan–Meier plot illustrating all-cause mortality in high-risk patients. (F) Pooled Kaplan–Meier plot illustrating all-cause mortality in intermediate-risk patients. (G) Pooled Kaplan–Meier plot illustrating all-cause mortality in low-risk patients. AVR, aortic valve replacement; IPD, individual patient data; SAVR, surgical aortic valve replacement; TAVR, transcatheter aortic valve replacement.

Correlation between STS PROM and 5-year mortality for AVR, regardless of TAVR or SAVR, is illustrated in Figure 2A and Supplementary Figures 4 and 5. Subgroup analyses based on STS PROM profiles were conducted to mitigate confounding due to varying risks. In the high-risk group (PARTNER 1 and CoreValve), no significant difference in all-cause mortality between TAVR and SAVR (P = 0.588) were observed over the 5-year follow-up (Figure 1D and 1E), with proportional hazards assumption violation (P < 0.001). TAVR exhibited lower all-cause mortality in the initial 6 months (HR 0.63, 95% CI: 0.48-0.82), but this benefit diminished after 6 months (HR 1.14, 95% CI: 0.97-1.34). In the intermediate-risk group (SAPIEN 3 and SURTAVI), comparable all-cause mortality was noted between TAVR and SAVR (P = 0.476), with non-proportional hazards (P = 0.046). TAVR showed lower risk in the first 6 months (HR 0.65, 95% CI: 0.48-0.88), with similar risks beyond 6 months (HR 1.05, 95% CI: 0.91-1.19) (Figure 1D and 1F). The low-risk group (Evolut, PARTNER 3, UK TAVI, and NOTION) exhibited no significant difference (P = 0.222) in all-cause mortality between TAVR and SAVR over the 5-year follow-up (HR 0.88, 95% CI: 0.71-1.10). In landmark analysis, TAVR demonstrated lower mortality risk up to 6 months (HR 0.54, 95% CI: 0.34-0.84) compared with SAVR, with similar risks after 6 months (HR 1.05, 95% CI: 0.81-1.36) (Figure 1D and 1G). Sensitivity analysis, excluding the cohort study SAPIEN 3 and only including RCTs, yielded similar results (Supplementary figure 3).

**Figure 2.**
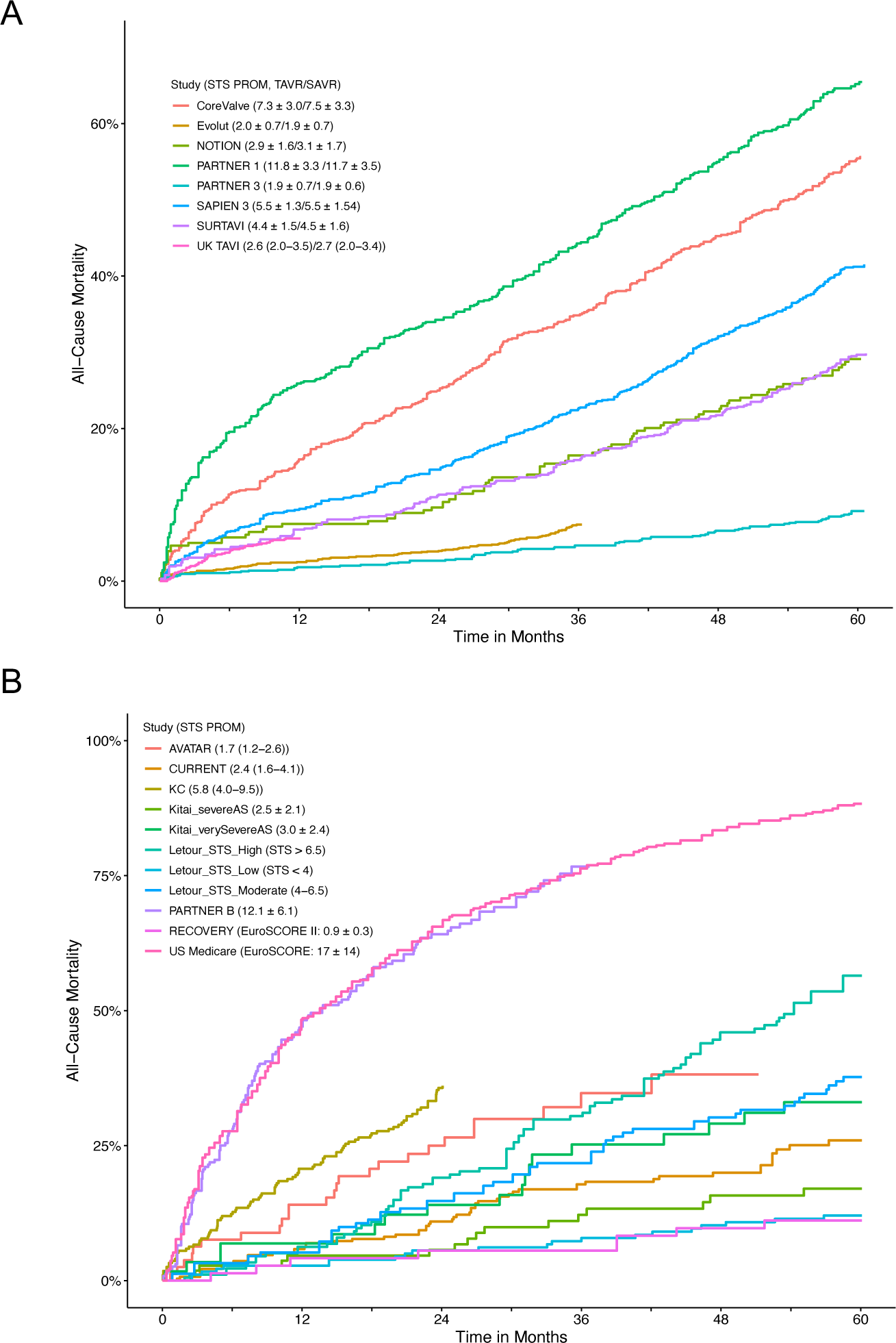
Correlation between STS PROM and all-cause mortality. (A) Kaplan-Meier plot of all-cause mortality from individual studies encompassing both TAVR and SAVR. (B) Kaplan–Meier plot displaying the relationship between STS PROM and all-cause mortality in individual studies, specifically focusing on conservative management with reported STS PROM or EuroSCORE. STS PROM, Society of Thoracic Surgeons Predicted Risk of Mortality; other abbreviation as in Figure 1.

### All-cause mortality in AVR versus conservative management

To investigate the benefits of AVR for treatment of severe AS, 10 RCTs (CoreValve, NOTION, Evolut, PARTNER 1, PARTNER 3, SURTAVI, UK TAVI, PARTNER B, AVATAR and RECOVERY) and 12 cohort studies (SAPIEN 3, Norway, KC registry, CURRENT, Letour_STS, US Medicare, Kitai, Korean, NEDA, Suzuki, California and Egnite) were included for meta-analysis. AVR significantly reduced all-cause mortality compared to conservative medical management (P < 0.001) with crude mortality rates at 5 years of 31.6% vs 49.3% (Figure 3A). The NNT was 5.7 at 5 years and the RMST was 8.9 months greater with AVR than conservative management (P < 0.001) (Figure 3D). Proportional hazards assumption violation was evident over the entire 60-month follow-up (P < 0.001). Flexible hazard regression analysis showed a time-varying HR favoring AVR over conservative medical management throughout the entire follow-up period (Figure 3C). Landmark analyses with a 40-month cutoff maintained the proportional hazards assumption, revealing a lower risk of mortality for AVR with HRs of 0.29 (95% CI: 0.25-0.33) up to 40 months and 0.28 (95% CI: 0.20-0.37) beyond 40 months (Figure 3B).

**Figure 3.**
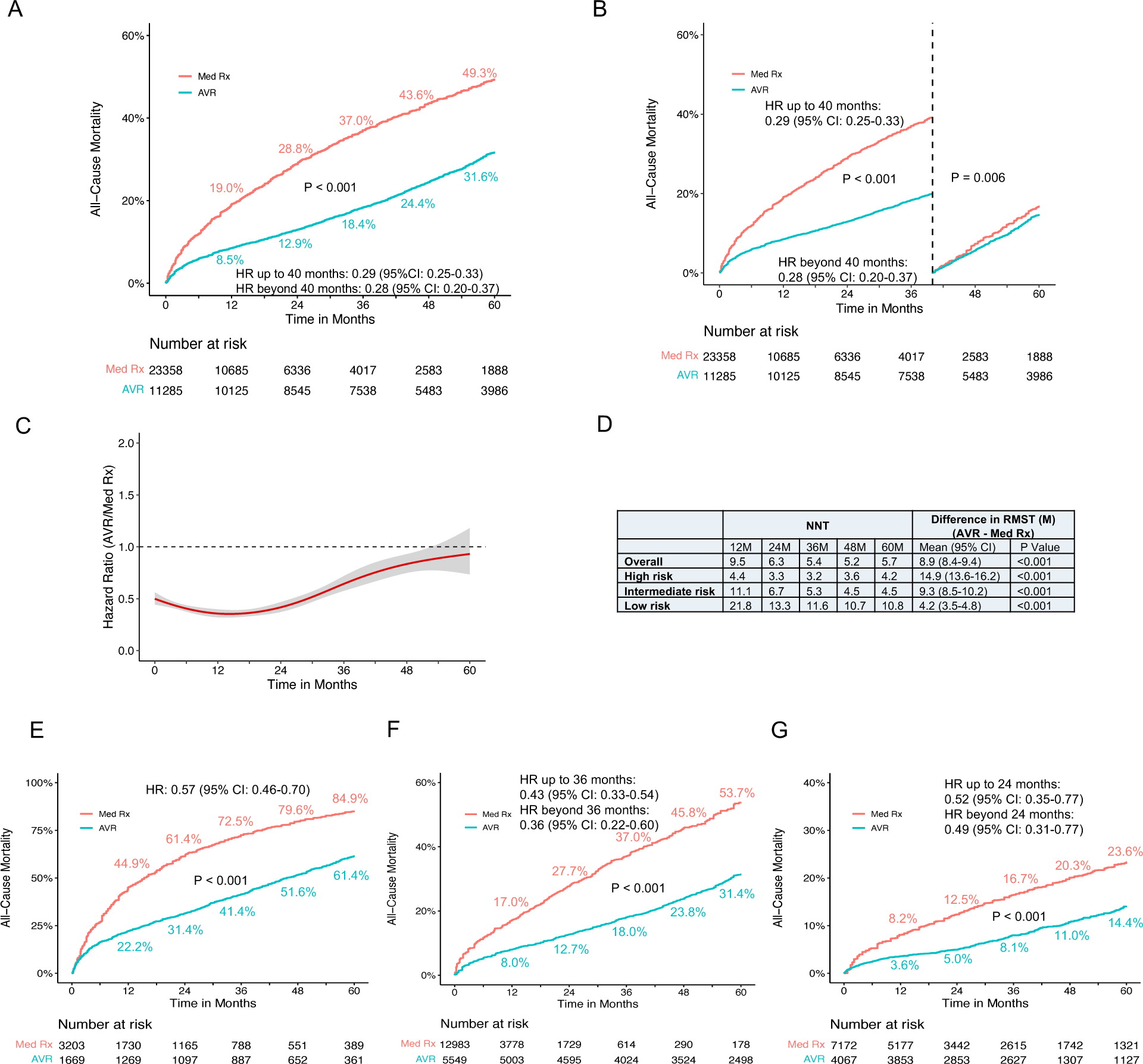
Time-to-Event Curves for all-cause mortality comparison between AVR and conservative management. (A) Pooled Kaplan–Meier plot depicting reconstructed IPD analysis for all-cause mortality post AVR, contrasting with medical conservative management. (B) Landmark analysis depicting all-cause mortality for AVR versus conservative management. (C) Hazard ratios with 95% confidence intervals illustrating the time-varying dynamics of all-cause mortality for AVR compared to medical conservative management. The red curved line represents the time-varying hazard ratios, and the grey area denotes the 95% confidence interval. (D) Number needed to treat (NNT) and the Difference in Restricted Mean Survival Time (RMST) are provided for the overall patient population and diverse risk subgroups. (E) Pooled Kaplan–Meier plot illustrating all-cause mortality in high-risk patients. (F) Pooled Kaplan–Meier plot illustrating all-cause mortality in intermediate-risk patients. (G) Pooled Kaplan–Meier plot illustrating all-cause mortality in low-risk patients. M, month; Med Rx, medical therapy; other abbreviation as in Figure 1.

Subgroup analyses, considering the inherent heterogeneity in cohort studies, were conducted based on different STS PROM or EuroSCORE scores. The correlation of STS PROM or EuroSCORE scores with long-term mortality for conservative medical therapy, except for AVATAR, was generally observed (Figure 2B and Supplementary Figure 6). In the high-risk group (PARTNER 1, CoreValve, US Medicare, Norway [conservative management arm], PARTNER B, California), crude mortality rates at 5 years were 61.4% for AVR and 84.9% for conservative management. The NNT was 4.2 at 5 years, and the RMST was 14.9 months greater with AVR (P < 0.001) (Figure 3D). There was a significant difference in all-cause mortality between AVR and conservative management (P < 0.001) during the 5-year follow-up with an HR of 0.57 (95% CI: 0.46-0.70) (Figure 3E). In the intermediate-risk group (SAPIEN 3, SURTAVI, KC registry, Suzuki, Korean, Letour_STS, Norway [AVR arm]), crude mortality rates at 5 years were 31.4% for AVR and 53.7% for conservative management. The NNT was 4.5 at 5 years, and the RMST was 9.3 months greater with AVR (P < 0.001) (Figure 3D). AVR was associated with a lower risk of all-cause mortality compared with conservative management (P < 0.001). Hazards changed non-proportionally over time in the intermediate-risk group (P = 0.040), with lower risk in AVR both in the first 36 months (HR 0.43, 95% CI: 0.33-0.54) and beyond 36 months (HR 0.36, 95% CI: 0.22-0.60) (Figure 3F).

In the low-risk group (Evolut, PARTNER 3, UK TAVI, NOTION, Kitai, CURRENT, NEDA, Letour_STS, RECOVERY, AVATAR), crude mortality rates at 5 years were 14.4% for AVR and 23.6% for conservative management. The NNT was 10.8 at 5 years, and the RMST was 4.2 months greater with AVR (P < 0.001) (Figure 3D). There was a significant lower risk of all-cause mortality for AVR throughout the 5-year follow-up (log-rank P < 0.001) (Figure 3F). Proportional hazards assumption violation was observed (P < 0.001). In the landmark analysis, AVR was associated with a lower risk of mortality both in the first 24 months (HR 0.52, 95% CI: 0.35-0.77) and after 24 months with an HR of 0.49 (95% CI: 0.31-0.77) compared to conservative management (Figure 3F). Sensitivity analysis, excluding the largest study Egnite or including only studies with reported STS PROM or EuroSCORE, yielded consistent results (Supplementary figures 7 and 8).

### Secondary endpoints

In the analysis of the composite of death from any cause, stroke or MI, seven RCTs (NOTION, Evolut, PARTNER 1, PARTNER 2, PARTNER 3, SURTAVI, and UK TAVI) and the TAVR arm from SAPIEN3 were included. No difference for the composite endpoint between TAVR and SAVR (P = 0.372) were observed (Figure 4A). There was also evidence of proportional hazards assumption violation in the entire follow-up period (P < 0.001). Landmark analyses with a cutoff of 6 months maintained the proportional hazards assumption, revealing a lower risk of composite endpoint for TAVR with an HR of 0.67 (95% CI: 0.58-0.79) in the first 6 months. Conversely, beyond 6 months, TAVR demonstrated a higher risk for composite endpoint (HR 1.20, 95% CI: 1.09-1.33) compared to SAVR (Supplementary figure 9A). This trend was consistently supported by flexible hazard regression analysis (Supplementary figure 9B). Sensitivity analysis, including only RCTs, produced similar results (Supplementary figure 9C).

**Figure 4.**
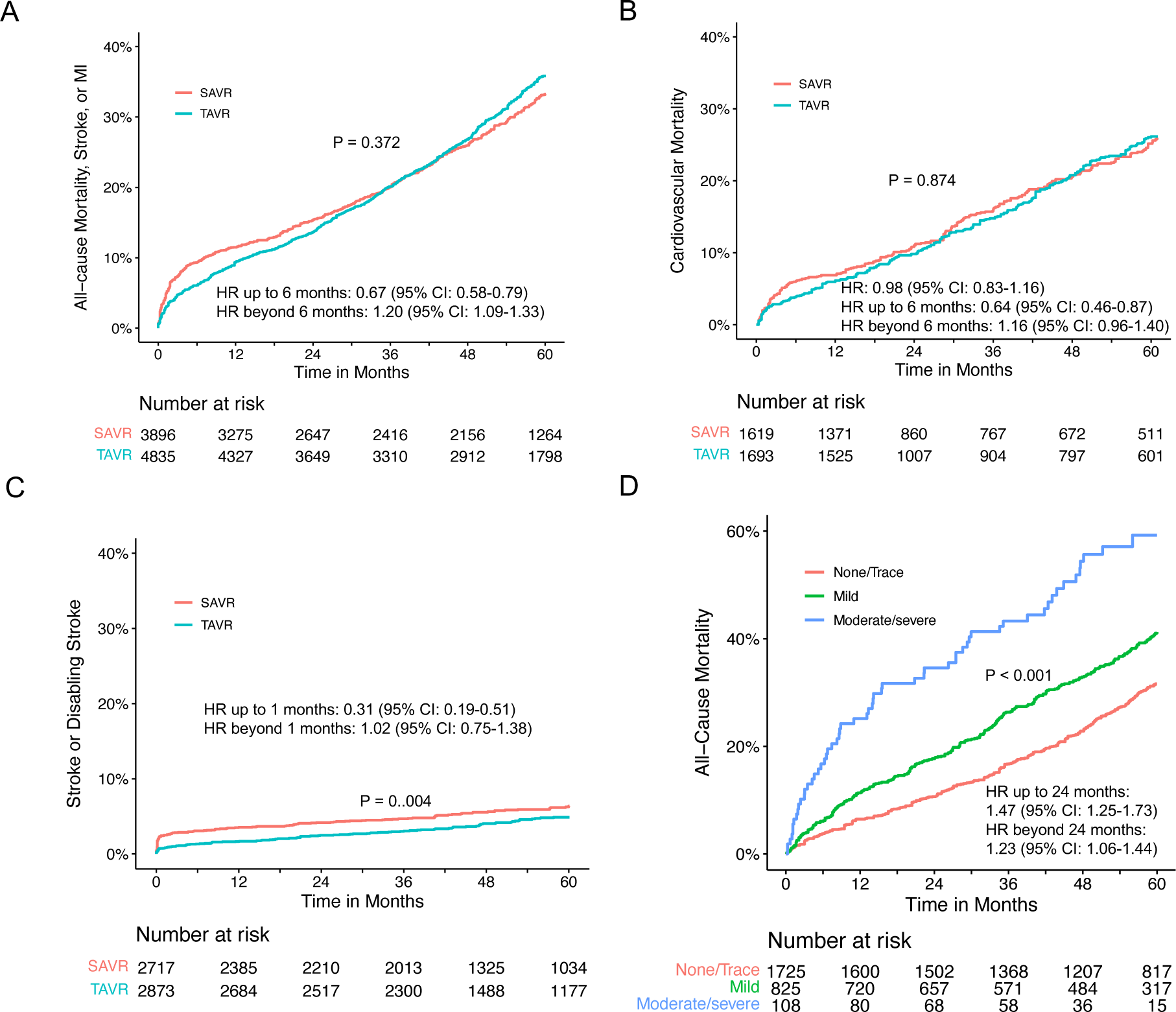
Secondary endpoints for TAVR and SAVR. Pooled Kaplan–Meier plot illustrating the composite endpoint of all-cause mortality, stroke, or MI (A), cardiovascular mortality (B), stroke or disabling stroke (C) following TAVR and SAVR. (D) Pooled Kaplan–Meier plot illustrating all-cause mortality stratified by the degree of paravalvular aortic regurgitation in patients undergoing TAVR. Abbreviation as in Figure 1.

For cardiovascular mortality, four RCTs (CoreValve, PARTNER 1, PARTNER 3, and UK TAVI) were included for meta-analysis. The incidence of cardiovascular mortality was comparable (P = 0.874) between TAVR and SAVR with an HR of 0.98 (95% CI: 0.83-1.16) (Figure 4B). Regarding stroke or disabling stroke, three studies (SURTAVI, SAPIEN3 and Evolut) reported the outcome of disabling stroke and 1 RCT (PARTNER 3) reported stroke. Pooled analysis revealed SAVR was associated with a higher rate of stroke or disabling stroke (P = 0.004). Landmark analysis revealed a high risk of stroke or disabling stroke in the first month (HR 0.31, 95% CI: 0.19-0.51), and a similar incidence of stroke after 1 month (HR 1.02, 95% CI: 0.75-1.38) (Figure 4C).

For all-cause mortality in patients with paravalvular regurgitation, a meta-analysis included five RCTs (PATNER 1, PATNER 2, PATNER 3, PATNER B and SURTAVI). A significant difference was observed among the various severity levels of paravalvular regurgitation (P < 0.001) (Figure 4D). The proportional hazards assumption did not hold during the follow-up period (P = 0.013). Landmark analysis revealed an increased risk of all-cause mortality in patients with increased severity levels of paravalvular regurgitation with an HR of 1.47 (95% CI: 1.25-1.73) in the first 24 months and an HR of 1.23 (95% CI: 1.06-1.44) beyond 24 months (Figure 4D).

### Hemodynamics

Six RCTs (Evolut, PARTNER 1, PARTNER 2, PARTNER 3, SURTAVI, and UK TAVI) provided data on mean aortic valve area and pressure gradient measured by serial echocardiograms pre- and post-operation. In addition, the SAPIEN3 cohort study, specifically from its TAVR arm, has contributed measurements of mean aortic valve area. Before intervention, the mean aortic valve effective orifice area were 0.74 cm^2^ (95% CI: 0.69-0.79) for TAVR and 0.76 cm^2^ (95% CI: 0.67-0.85) for SAVR. At one month post-operatively, mean aortic valve effective orifice area for TAVR and SAVR were 1.84 cm^2^ (95% CI: 1.47-2.20) and 1.71 cm^2^ (95% CI: 1.43-1.99), respectively. The mean effective orifice area appeared relatively stable over the 5-year follow-up period (Figure 5A). Pre-operative mean aortic valve pressure gradients were 45.6 mmHg (95% CI: 43.6-47.6) for TAVR and 45.5 mmHg (95% CI: 43.3-47.6) for SAVR. At one month, these values decreased to 10.0 mmHg (95% CI: 7.9-12.2) and 11.0 mmHg (95% CI: 9.9-12.1), respectively. Similarly, the mean pressure gradient remained consistent throughout the 5-year follow-up period (Figure 5B).

**Figure 5.**
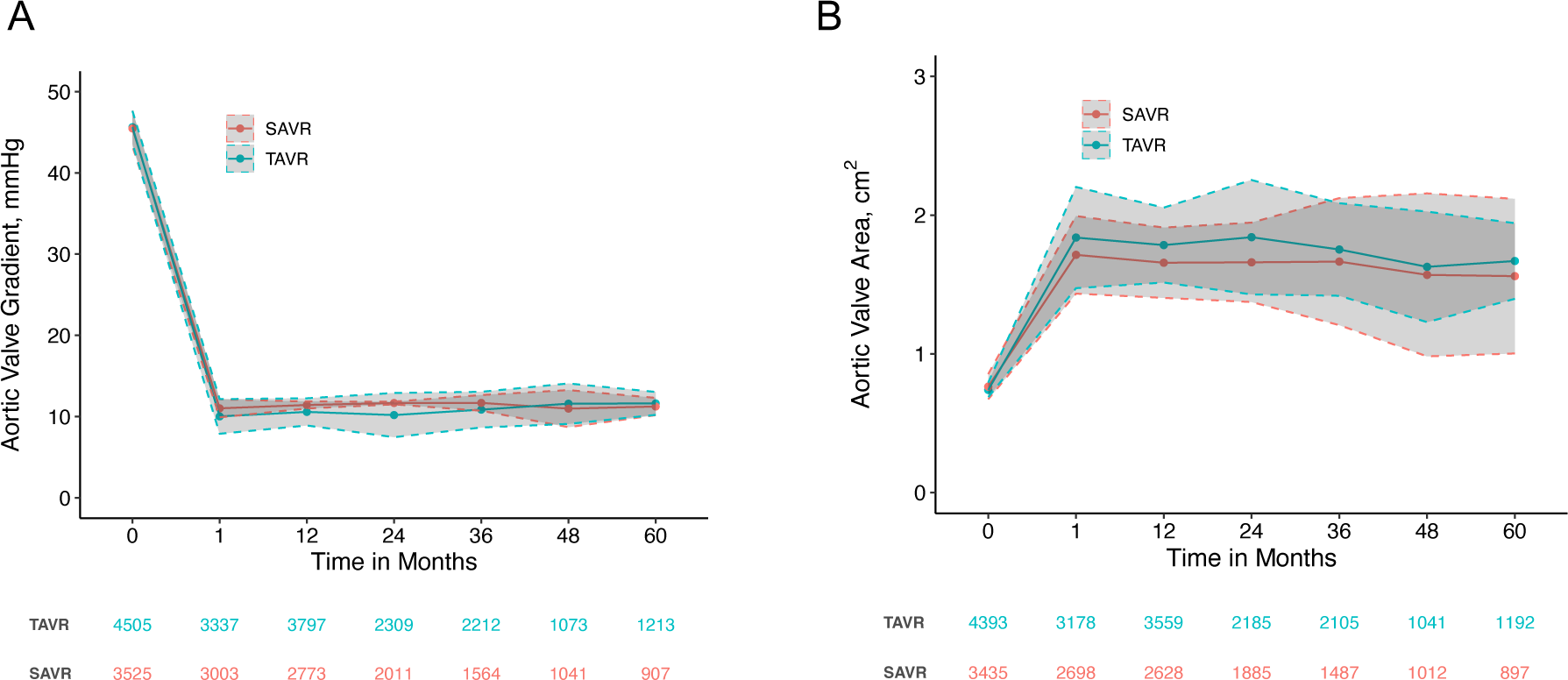
Long-term hemodynamic meta-analysis of aortic valve gradient and orifice area in transcatheter versus surgical patients over 5 years. (A) Weighted mean with solid lines and 95% confidence intervals with dotted lines for aortic valve pressure gradient. (B) Weighted mean with solid lines and 95% confidence intervals with dotted lines for aortic valve orifice area.

## Discussion

Aortic stenosis, a progressive disease, poses a substantial public health concern in elderly, marked by significant mortality and morbidity ^3^. SAVR has long been the cornerstone for treatment of symptomatic severe AS. Over the last decade, TAVR has emerged as a viable alternative, even for patients at low risk of operative mortality. However, determining the superior treatment modality for severe AS remains inconclusive. In this meta-analysis involving patients with severe AS and utilizing Kaplan-Meier–derived IPD from RCTs, we observed no significant difference in all-cause mortality between TAVR and SAVR over the 5-year follow-up period, irrespective of operative mortality risk profiles. Notably, TAVR exhibited a significantly lower risk of all-cause death in the initial 6 months. Additionally, by synthesizing data from both RCTs and cohort studies, our analysis revealed a notable association between AVR and lower risk for all-cause death.

Evidence has consistently shown that STS PROM score is a robust predictor. It has been documented that STS PROM not only forecasts 30-day mortality but also extends its predictive accuracy to long-term mortality after cardiac surgical procedures ^43^. In alignment with this, our study illustrates a robust correlation between STS PROM and 5-year mortality for patients undergoing SAVR, as well as those opting for TAVR. Additionally, EuroSCORE II, a metric comparable to STS PROM in predicting operative mortality, demonstrates superior accuracy compared to EuroSCORE I, which tends to overestimate operative mortality ^44^. Based on these, we present the first demonstration that STS PROM and EuroSCORE can also effectively predict 5-year mortality in severe AS patients undergoing conservative management. Building upon these insights, we proposed that these risk scores can be extrapolated from survival curves in studies lacking reported STS PROM, facilitating the categorization of patients into subgroups to mitigate heterogeneities and biases. Our sensitivity analyses validated the robustness and consistency of these findings across studies, regardless of STS PROM reporting status.

Aligned with previously published data, the proportional hazards assumptions for all-cause mortality in overall, high, and intermediate-risk groups were found to be violated. Employing STS PROM, our analysis revealed no significant differences in all-cause mortality between TAVR and SAVR in high and intermediate-risk groups over the follow-up period. Notably, TAVR exhibited superiority in the initial 6 months, corroborating findings reported in high-risk group, including both high and intermediate-risk individuals, stratified according to STS PROM ^45^. In the low-risk group, leveraging extended follow-up data compared to previous studies, our study yielded consistent results like those observed in the high and intermediate-risk groups. It is worth noting that a recent meta-analysis of low-risk trials, combined with propensity score-matched registry data, suggested higher mortality with TAVR after 2 years compared to SAVR ^46^. However, this difference was driven by the propensity score-matched registry data. Considering the limited data on the mortality burden for severe AS in the modern era and the associated mortality benefits after AVR, our systematic analysis robustly demonstrated a significant association between AVR and mortality benefits. This effect was particularly pronounced in high and intermediate-risk groups, characterized by a low NNT and prolonged survival gain. Even in low-risk groups, encompassing asymptomatic severe AS cases, our synthesized data advocates for AVR over conservative management for mortality benefits. Ongoing clinical trials such as EARLY TAVR ^47^ and EVOLVED ^48^ are anticipated to provide further insights into the role of early TAVR in asymptomatic severe AS compared to conventional clinical surveillance.

Our study has several inherent limitations. Firstly, the inclusion of data synthesized from different generations of transaortic valves, employing varied deployment strategies, may not fully capture the current landscape of clinical practice. Secondly, the use of IPD derived from Kaplan–Meier plots, while considered a well-accepted approach, may introduce some compromise in data quality compared to a true IPD meta-analysis. Additionally, there is a recognized risk of bias in RCTs comparing TAVR and SAVR ^49^. Thirdly, the diverse outcomes reported across trials, coupled with variations in definitions and follow-up durations, could introduce selection and measurement bias when amalgamating data for meta-analysis. Although we attempted to address study-level heterogeneity through appropriate modeling, the potential for bias persists. Fourthly, the paucity of randomized clinical trials directly comparing AVR to conservative medical management led to the inclusion of many cohort studies. Despite efforts to mitigate confounding factors by stratification using STS PROM, the retrospective nature of these studies introduces inherent limitations, and residual confounders and bias may persist. Lastly, the generalizability of our findings may be limited, as they might not be readily extrapolated to patient populations excluded from the trials, such as those with bicuspid aortic valves, preexisting bioprosthetic or mechanical heart valves, and younger individuals. These exclusions underscore the importance of interpreting our results within the specified study population parameters.

In summary, this meta-analysis, utilizing Kaplan-Meier–derived IPD, offers valuable insights into the comparative outcomes of TAVR compared to SAVR and conservative management compared with AVR, for severe AS. Our findings reveal no significant disparities in all-cause mortality and cardiovascular mortality over a 5-year period between TAVR and SAVR. However, TAVR demonstrated a noteworthy advantage with a notably lower risk of all-cause and cardiovascular mortality in the initial 6 months. Furthermore, TAVR exhibited a reduced risk of stroke compared to SAVR during the first month post-procedure. The mean aortic valve area and pressure gradient remained comparable between TAVR and SAVR, exhibiting stability throughout the 5-year follow-up. Additionally, AVR was associated with a considerable mortality benefit when compared to conservative management, regardless of risk profile. This comprehensive analysis not only contributes to our understanding of mortality outcomes for severe AS patients across different risk categories and treatment approaches but also holds significant implications for informing clinical decision-making and shaping policies in the field of structural cardiology.

## Data Availability

All data produced in the present work are contained in the manuscript

## Notes

### Competing Interest Statement

The authors have declared no competing interest.

### Clinical Protocols

https://www.crd.york.ac.uk/prospero/display_record.php?RecordID=508950

### Funding Statement

This study did not receive any funding

### Author Declarations

Source data were openly available.

