## Supplemental data for "Transcatheter versus Surgical Interventions for Severe Aortic Stenosis: A Contemporary Evaluation against Conservative Management"

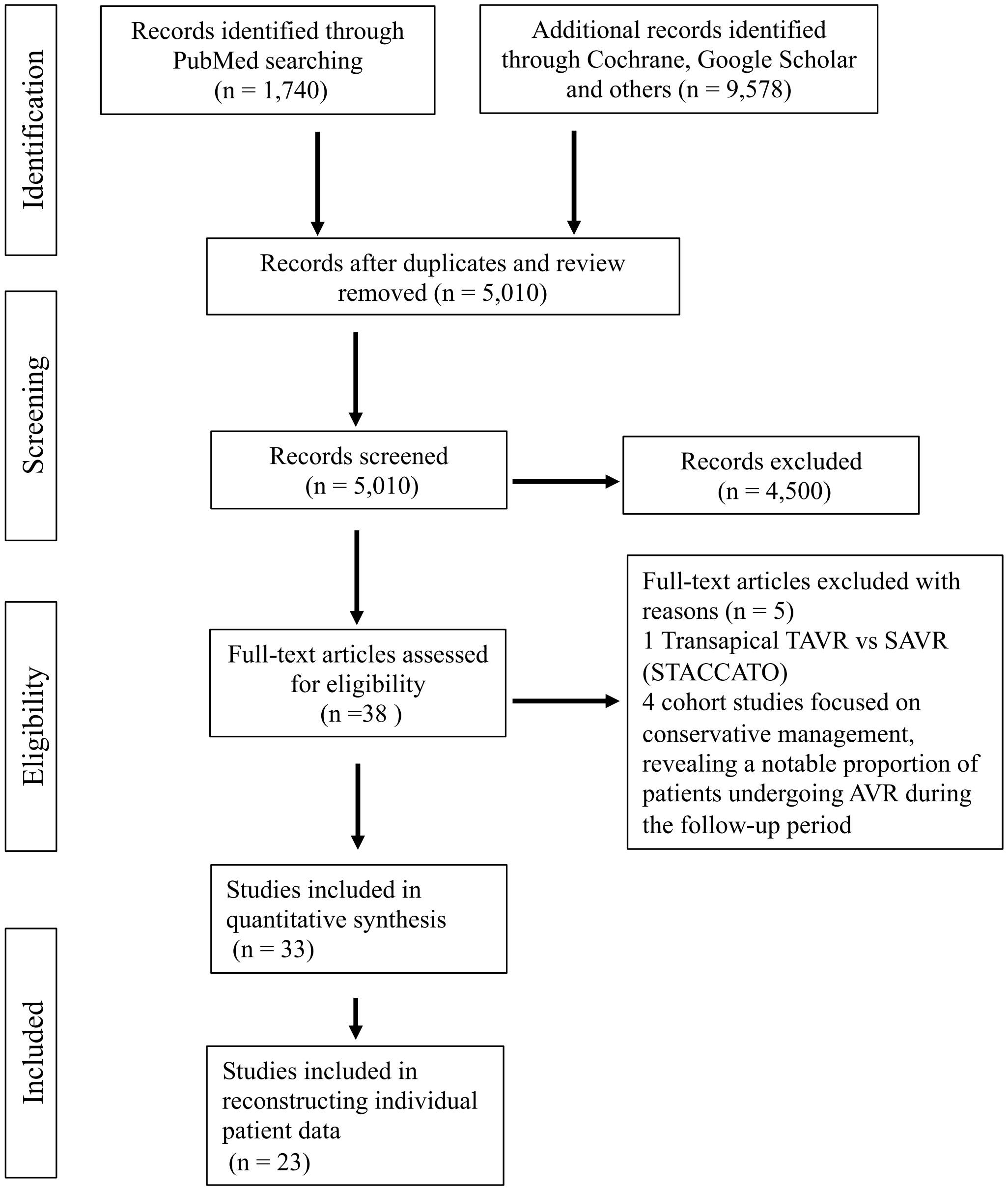


Supplementary figure 1. Flowchart illustration of the study selection.

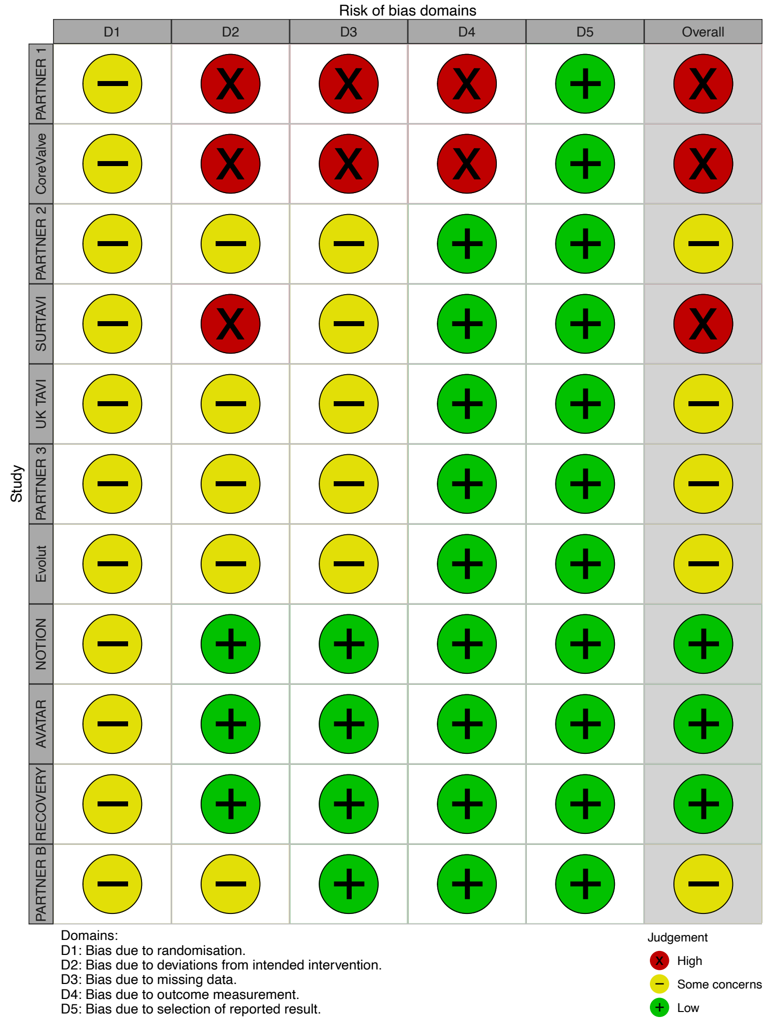


Supplementary figure 2. Risk of Bias Assessment for randomized trials.

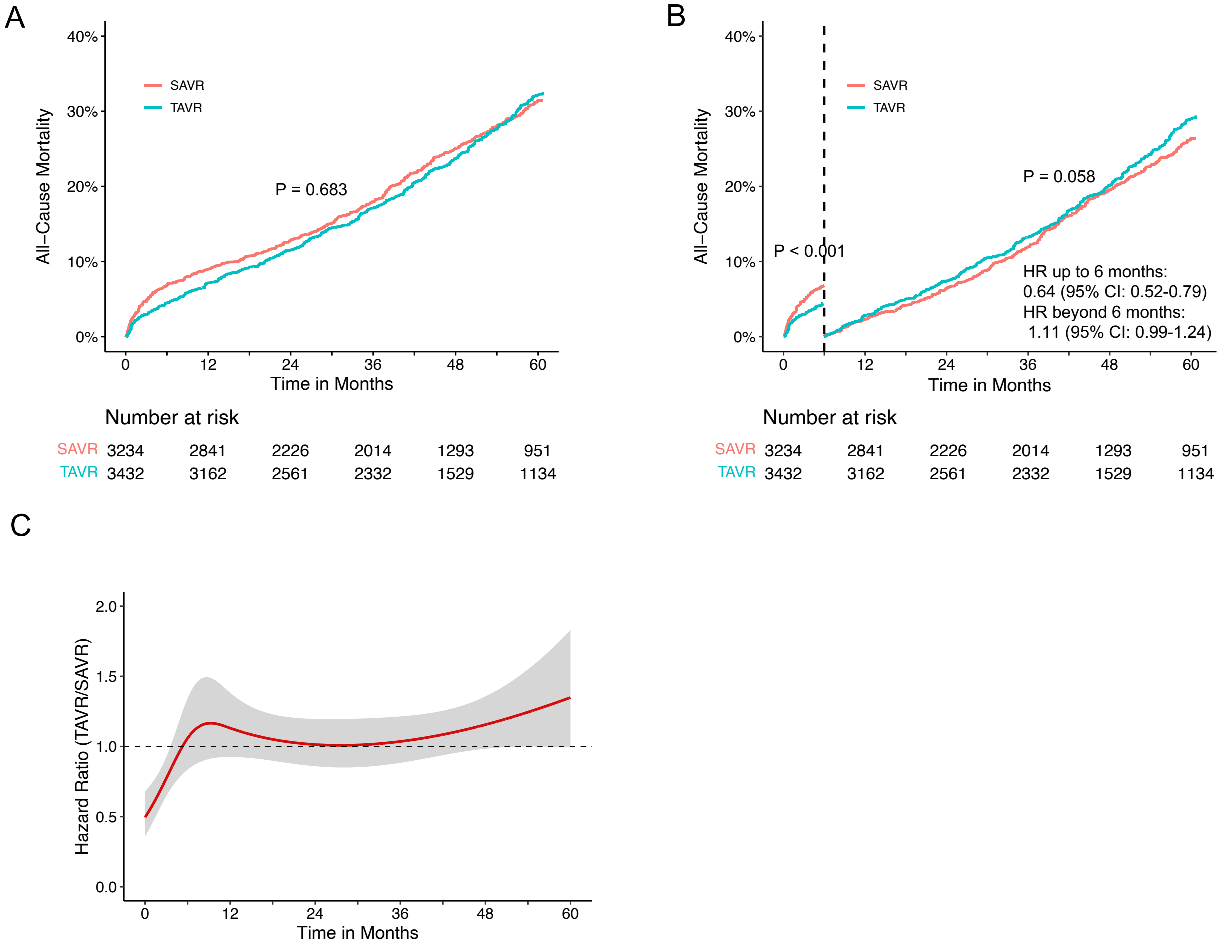


Supplementary figure 3. All-cause mortality for TAVR and SAVR in randomized trials. (A) Pooled Kaplan–Meier plots illustrating all-cause mortality in randomized trials, excluding SAPIEN3 from Figure 1A. (B) Landmark analysis of all-cause mortality, comparing TAVR versus SAVR specifically within randomized trials. (C) Time-varying Hazard Ratios with 95% Confidence Intervals, delineating the dynamic trends in all-cause mortality for TAVR versus SAVR within randomized trials. SAVR, surgical aortic valve replacement; TAVR, transcatheter aortic valve replacement.

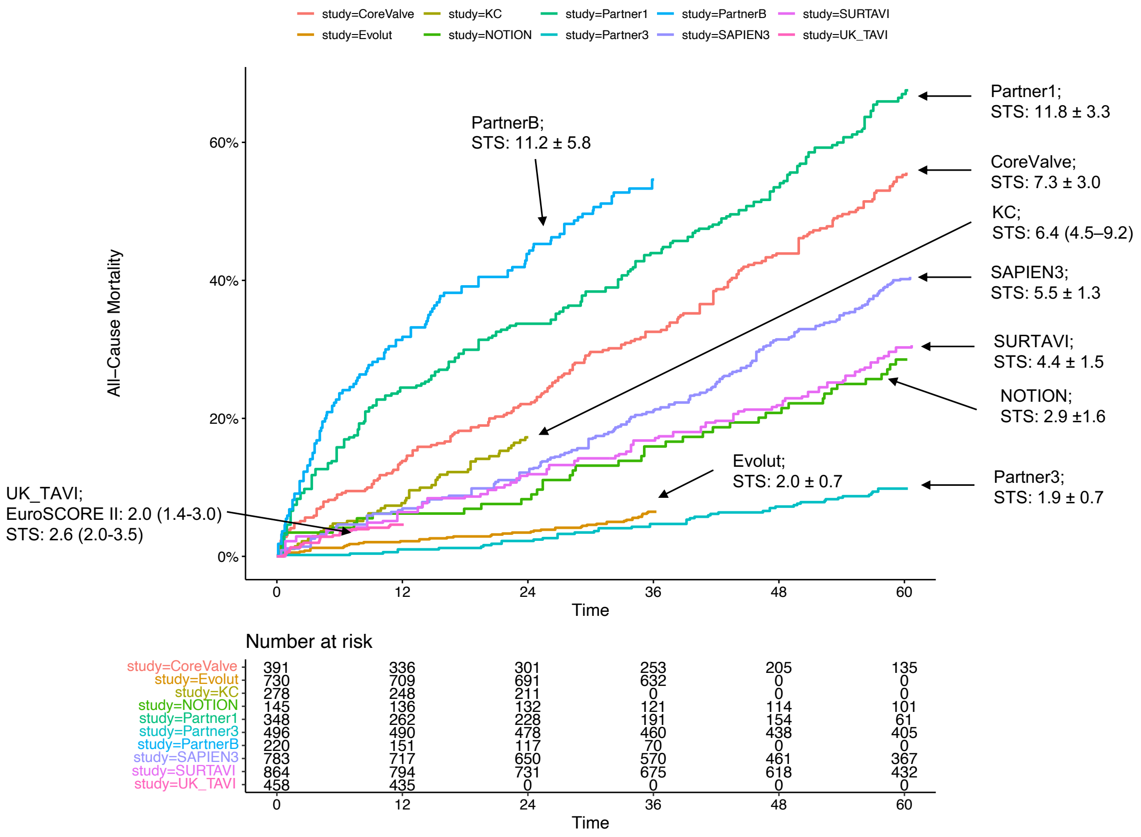


Supplementary figure 4. Robust correlation between the Society of Thoracic Surgeons Predicted Risk of Mortality (STS PROM) and all-cause mortality in patients undergoing TAVR. Each data point on the plot is labeled with its corresponding STS PROM if reported, emphasizing the alignment between mortality outcomes and the predicted risk scores. TAVR, transcatheter aortic valve replacement.

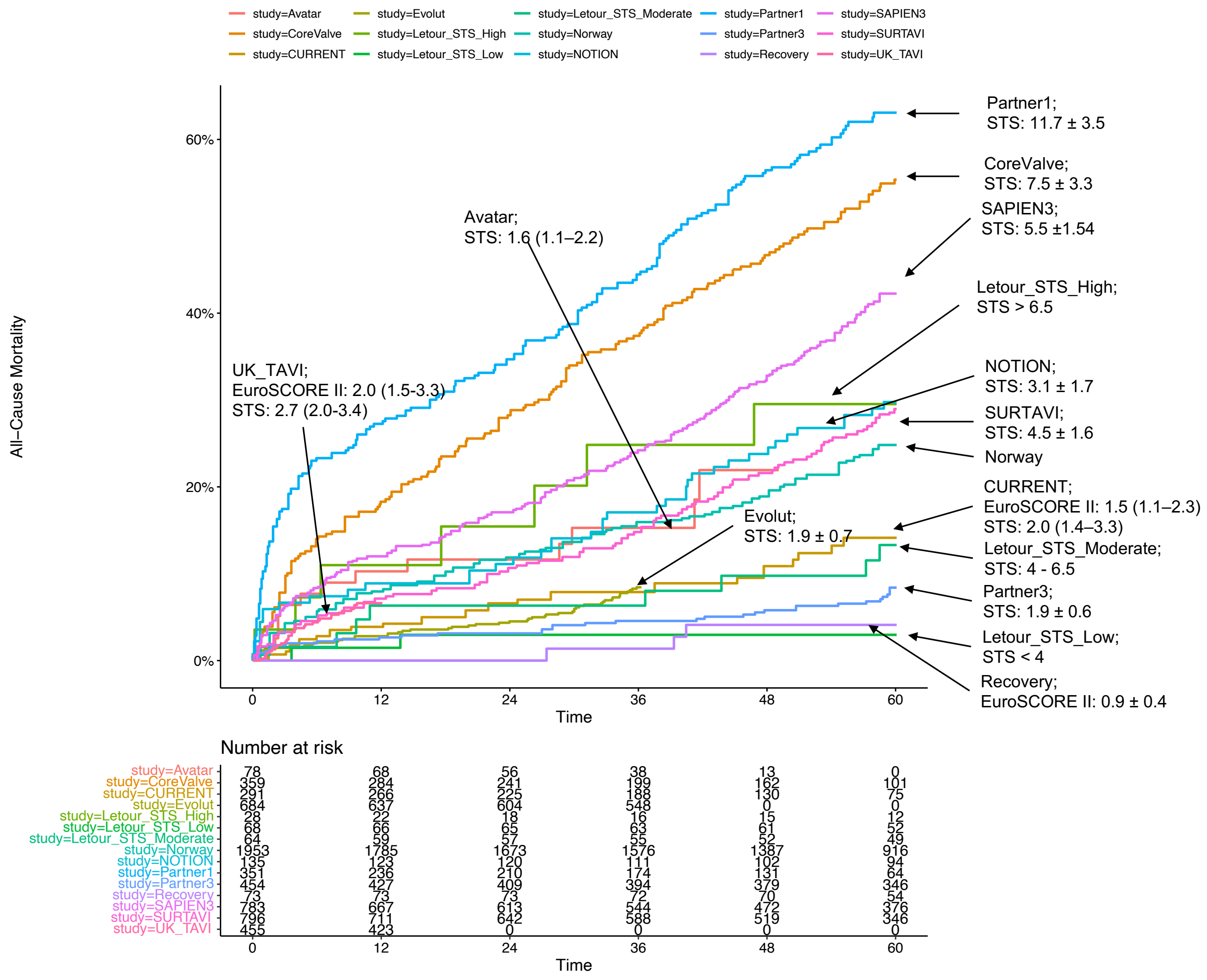


Supplementary figure 5. Robust correlation between the Society of Thoracic Surgeons Predicted Risk of Mortality (STS PROM) and all-cause mortality in patients undergoing SAVR. Each data point on the plot is labeled with its corresponding STS PROM if reported, emphasizing the alignment between mortality outcomes and the predicted risk scores. SAVR, surgical aortic valve replacement. STS, Society of Thoracic Surgeons Predicted Risk of Mortality.

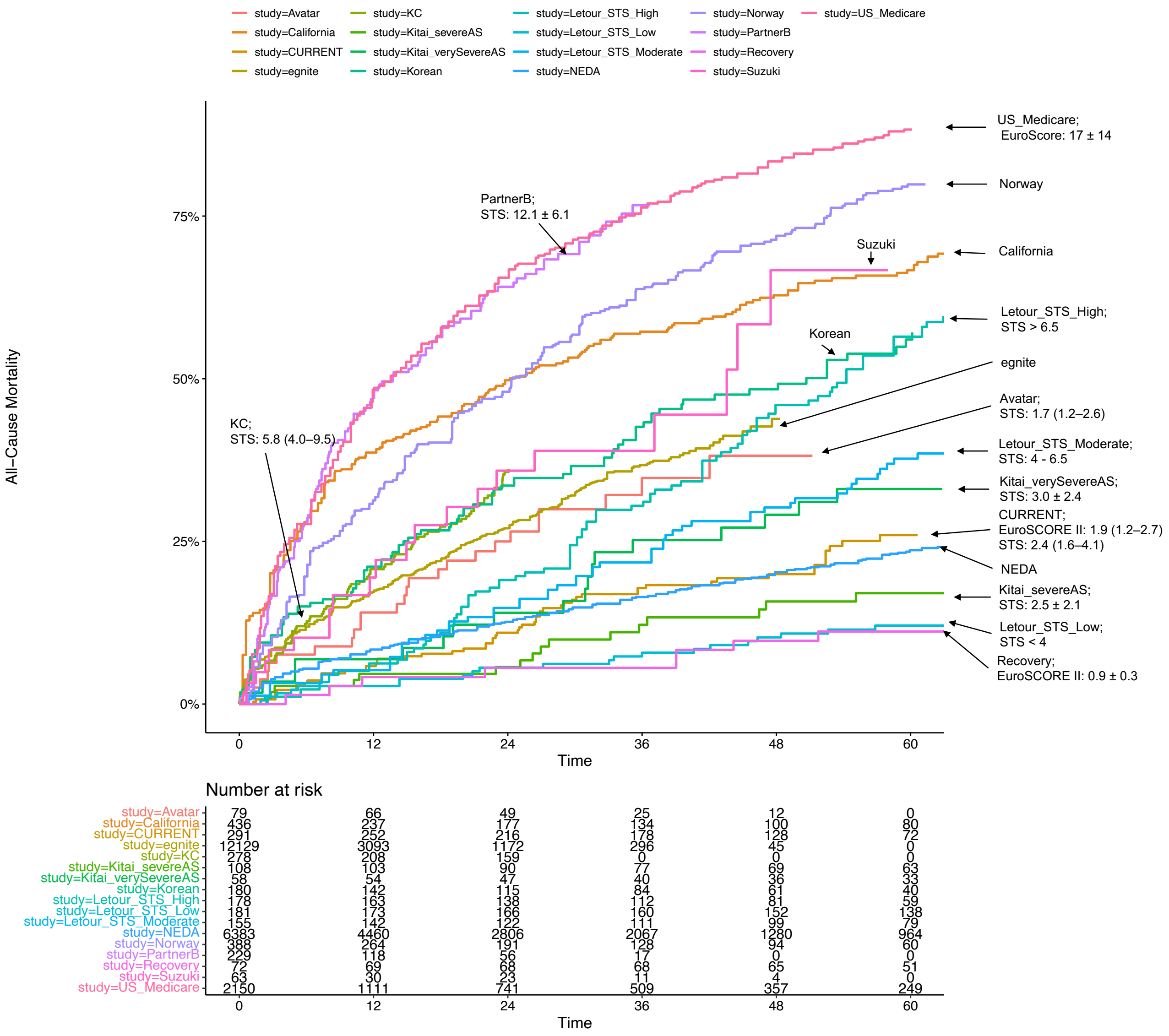


Supplementary figure 6. Correlation between the Society of Thoracic Surgeons Predicted Risk of Mortality (STS PROM) and all-cause mortality in patients undergoing conservative management. Each data point on the plot is labeled with its corresponding STS PROM if reported, emphasizing the alignment between mortality outcomes and the predicted risk scores.

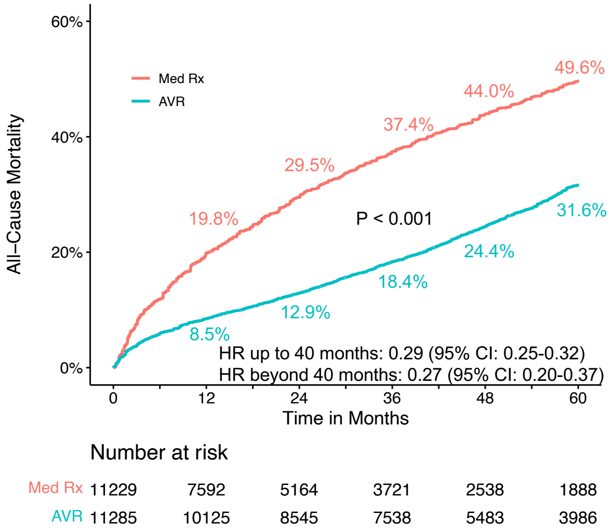


Supplementary figure 7. Sensitivity analysis of all-cause mortality for AVR versus medical conservative management excluding the Egnite study from Figure 3C. AVR, aortic valve replacement; Med Rx, medical therapy.

A

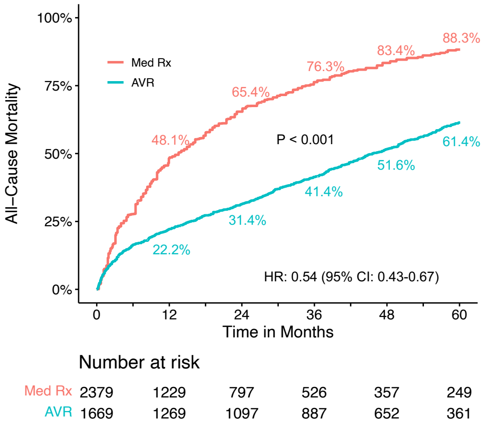


B

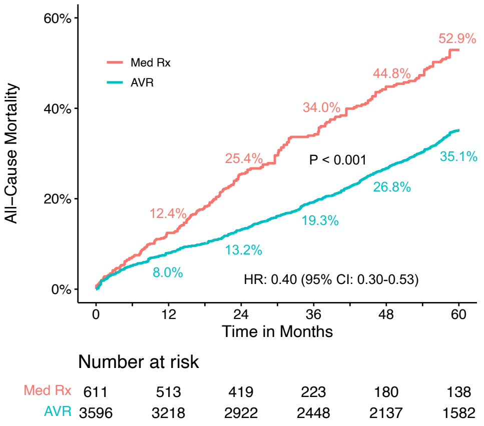


C

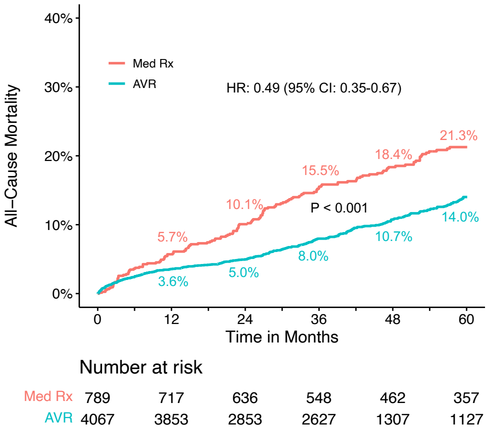


D

| | NNT | | | | | Difference in RMST (M) (AVR - Med Rx) | |
|---|---|---|---|---|---|---|---|
| | 12M | 24M | 36M | 48M | 60M | Mean (95% CI) | P Value |
| **High risk** | 3.8 | 2.9 | 32.9 | 3.1 | 3.7 | 16.8 (15.5-18.2) | <0.001 |
| **Intermediate risk** | 22.7 | 8.2 | 6.8 | 5.5 | 5.6 | 7.0 (5.1-8.9) | <0.001 |
| **Low risk** | 46.7 | 19.6 | 13.2 | 13.1 | 13.8 | 3.1 (1.9-4.4) | <0.001 |

Supplementary figure 8. Sensitivity analysis of all-cause mortality for AVR versus medical conservative management in studies with reported STS PROM. (A) Pooled Kaplan–Meier plot for all-cause mortality in high-risk patients. (B) Pooled Kaplan–Meier plot for all-cause mortality in intermediate-risk patients. (C) Pooled Kaplan–Meier plot for all-cause mortality in low-risk patients. (D) Number Needed to Treat (NNT) and the Difference in Restricted Mean Survival Time (RMST) across these different risk groups. AVR, aortic valve replacement; Med Rx, medical therapy; STS PROM, Society of Thoracic Surgeons Predicted Risk of Mortality.

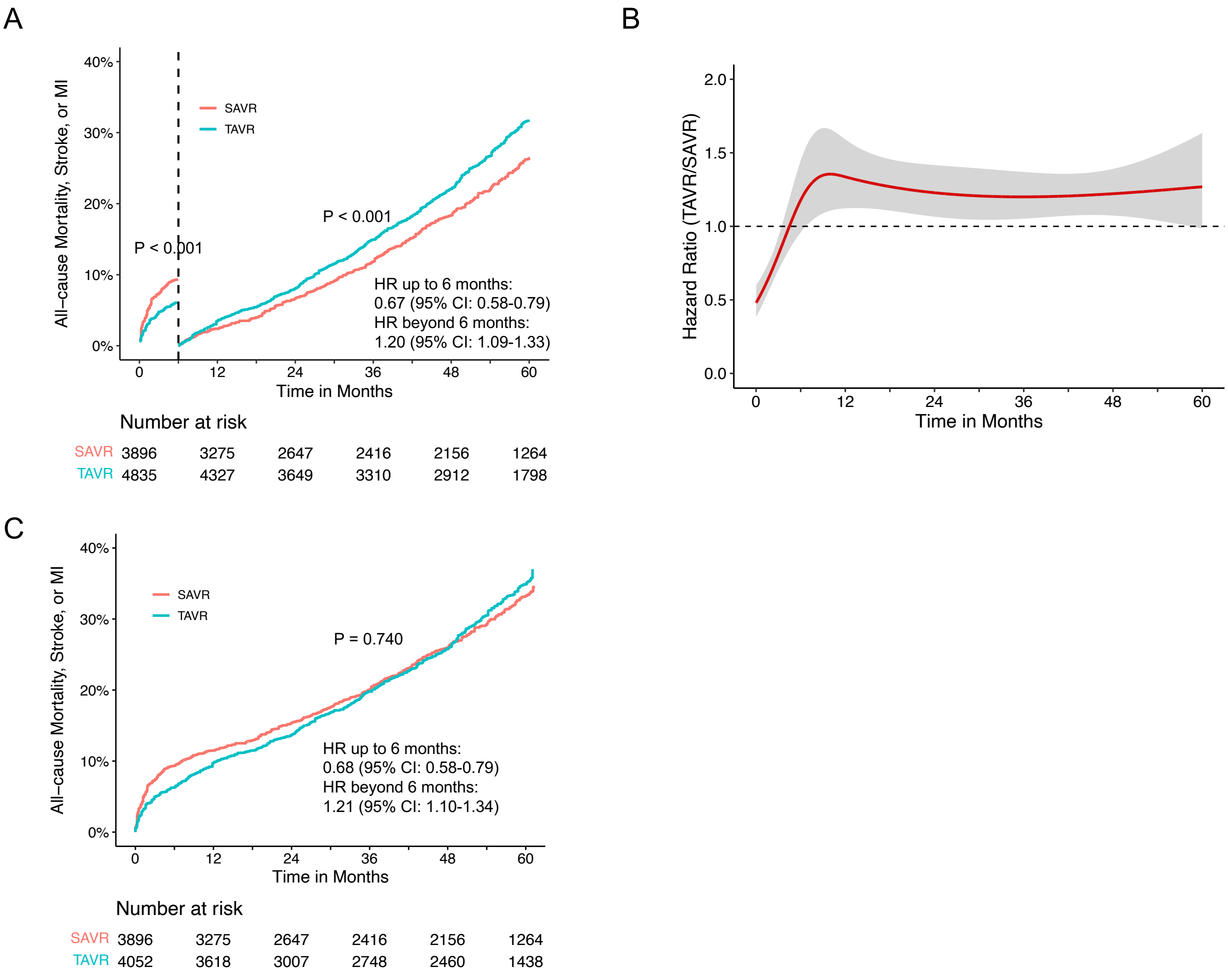


Supplementary figure 9. The composite of all-cause mortality, stroke or MI for TAVR versus SAVR. (A) Landmark analysis of the composite of all-cause mortality, stroke or MI for TAVR versus SAVR. (B) Time-varying hazard ratios with 95% CIs for the composite of all-cause mortality, stroke or MI for TAVR versus SAVR in randomized trials. (C) Pooled Kaplan–Meier plots for the composite endpoint in randomized trials, excluding SAPIEN3. MI, myocardial infarction; SAVR, surgical aortic valve replacement; TAVR, transcatheter aortic valve replacement.

Supplementary table 1. Electronic Database Search Strategy (As of 12/7/2023).

| Database | Search Strategy | Search Results |
|---|---|---|
| PubMed | ("Transcatheter aortic valve replacement"[Title/Abstract] OR "transcatheter aortic valve implantation"[Title/Abstract]) OR TAVR[Title/Abstract]) OR TAVI[Title/Abstract] OR "surgical aortic valve replacement"[Title/Abstract] OR "surgical aortic valve replacement"[Other Term] OR "SAVR"[Title/Abstract]) AND ("outcome*"[Title/Abstract] OR "follow-up"[Title/Abstract]) AND ((“controlled study”[Publication Type] OR “multicenter study”[Publication Type] OR "randomized controlled trial"[Publication Type] OR "trial"[Title/Abstract]) | 1,504 |
| | (“Severe Aortic Stenosis”[Title/Abstract]) AND ("Medical therapy”[Title/Abstract] OR "Conservative management"[Title/Abstract] OR "Natural history"[Title/Abstract] OR "Natural course"[Title/Abstract]) | 236 |
| Google Scholar | ("Transcatheter aortic valve replacement" OR "transcatheter aortic valve implantation" OR “TAVR” OR “TAVI” OR "surgical aortic valve replacement" OR "SAVR") ANA (“Severe Aortic Stenosis”) AND ("outcome" OR "follow-up") AND (“controlled study” OR “multicenter study” OR “randomized controlled trial”) | 3,410 |
| | (“Severe Aortic Stenosis”) AND ("Medical therapy” OR "Conservative management" OR "Natural history" OR "Natural course") AND ("outcome" OR "follow-up") AND (“All-cause mortality”) | 4,720 |
| Cochrane | ("Transcatheter aortic valve replacement" OR "transcatheter aortic valve implantation" OR “TAVR” OR “TAVI” OR "surgical aortic valve replacement" OR "SAVR") | 1,428 |
| | (“Severe Aortic Stenosis”) AND ("Medical therapy” OR "Conservative management" OR "Natural history" OR "Natural course") | 20 |

Supplementary table 2. Characteristics of the included trials and studies.

| Study | Study acronym | Study design | Region | N | Mean age | Follow up (Year) | Key Criteria | Intervention | Primary endpoint | Risk of bias |
|---|---|---|---|---|---|---|---|---|---|---|
| Adams et al. 2014; Gleason et al. 2018 | CoreValve | RCT | USA | 797 | 83.2 | 5 | Severe aortic stenosis and heart-failure symptoms of New York Heart Association (NYHA) class II or higher. Aortic stenosis was defined as AVA ≤0.8 $cm^2$ (index 0.5 $cm^2/m^2$) or peak velocity >4 m/s or mean PG >40 mmHg. High surgical risk was defined as an estimated 30-day risk of surgical mortality and major morbidity of at least 15%, but <50%. | TAVR vs SAVR | All-cause mortality at 1 year | ROB2: High |
| Popma et al. 2019; Forrest et al. 2023a; Forrest et al. 2023b; | Evolut | RCT | Australia, Canada, France, Japan, the Netherla-nds, New Zealand, and USA | 1414 | 74 | 3 and 4 | Severe aortic valve stenosis with trileaflet aortic valve morphology and a low predicted risk of death (<3%) from surgery. Severe, symptomatic AS AVA ≤ 1.0 $cm^2$ (index 0.6 $cm^2/m^2$) with peak velocity ≥ 4 m/s or mean PG ≥ 40 mmHg. | TAVR vs SAVR | Composite of all-cause mortality or disabling stroke at 2 years | ROB2: Some concerns |
| Thyregod et al. 2019; Jørgens-en et al. 2021 | NOTION | RCT | Denmark, Sweden | 280 | 79.1 | 8 | Severe, symptomatic AS AVA ≤1 m2 (index 0.6 $cm^2/m^2$) and either peak velocity >4 m/s or mean PG >40 mmHg. Patients with acute treatment, severe coronary artery disease, severe nonaortic valvular disease, prior heart surgery, recent stroke or myocardial infarction (MI), or severe lung or renal disease were excluded. | TAVR vs SAVR | Composite rate of all-cause mortality, stroke, or MI at 1 year and 5 years | ROB2: Low |
| Smith et al. 2011; Mack et al. 2015 | PARTNER 1 | RCT | USA, Canada, Germany | 699 | 84.1 | 5 | Severe, symptomatic AS AVA ≤0.8 cm2 (index 0.5 $cm^2/m^2$) or peak velocity ≥4 m/s or mean PG ≥40 mmHg. Patients were deemed to be at high risk for operative complications or death with an STS risk score of at least 10%. | TAVR vs SAVR | All-cause mortality at 1 year | ROB2: High |
| Leon et al. 2016; Makkar et al. 2020 | PARTNER 2 | RCT | USA, Canada | 2032 | 81.6 | 5 | Severe, symptomatic AS AVA ≤0.8 cm2 (index 0.5 $cm^2/m^2$) or peak velocity >4 m/s or mean PG >40 mmHg. Patients were deemed to be at intermediate risk for operative complications or death with an STS risk score of 4-8%. | TAVR vs SAVR | Composite of death from any cause or disabling stroke at 2 years | ROB2: Some concerns |
| Mack et al. 2019; Leon et al. 2021; Mack et al. 2023 | PARTNER 3 | RCT | USA, Australia, New Zealand, Japan | 1000 | 73.3 | 5 | Severe, symptomatic AS AVA ≤1.0 cm2 (index 0.6 $cm^2/m^2$) with peak velocity ≥4 m/s or mean PG ≥40 mmHg. Asymptomatic if LVEF <50% or abnormal exercise | TAVR vs SAVR | Composite of death from any cause, stroke, or rehospitalizat ion at 1 year | ROB2: Some concerns |

| | | | | | | | | | | |
|---|---|---|---|---|---|---|---|---|---|---|
| | | | | | | | test. Patients were deemed to be at intermediate risk for operative complications or death with an STS risk score of < 4%. | | | |
| Reardon et al. 2017; Van Mieghem et al. 2022 | SURTAVI | RCT | USA, Canada, Germany, The Netherla-nds, UK, Spain, Switzerla-nd, Sweden, Denmark | 1746 | 79.8 | 5 | Severe, symptomatic AS AVA ≤1.0 cm2 (index 0.6 $cm^2/m^2$) with peak velocity >4 m/s or mean PG >40 mmHg or Doppler velocity index <0.25. Patients were deemed to be at intermediate risk for operative complications or death with an STS risk score of 3-15%. | TAVR vs SAVR | Composite of death from any cause or disabling stroke at 2 years | ROB2: High |
| Toff et al. 2022 | UK TAVI | RCT | UK | 913 | 81 | 1 | Severe, symptomatic AS Age ≥80 or ≥70 with intermediate or high-risk. | TAVR vs SAVR | All-cause mortality at 1 year | ROB2: Some concerns |
| Banovic et al. 2022 | AVATAR | RCT | Belgium, Czech Republic, Italy, Croatia, Lithuania, Poland, and Serbia | 151 | 67 | 2 | Patients >18 years old presenting with severe AS. Patients were excluded if they had exertional dyspnea, syncope or presyncope, angina, a LVEF <50%, severe AS (defined as maximal aortic jet velocity >5.5 m/s at rest), aortic regurgitation ≥3+, dilatation of the ascending aorta requiring replacement of aortic root or ascending aorta (>5 cm), or significant mitral valve disease, or if they had undergone previous cardiac surgery. | SAVR vs Med Rx | All-cause mortality | ROB2: Low |
| Kang et al. 2020 | RECOVERY | RCT | Korea | 145 | 64 | 8 | Patients who were 20 to 80 years of age and who presented with very severe aortic stenosis (an aortic-valve area of 0.75 $cm^2$ or less with either a peak aortic jet velocity of 4.5 m per second or greater or a mean transaortic gradient of 50 mm Hg or greater). Patients were excluded if they had exertional dyspnea, syncope, presyncope or angina, a left ventricular ejection fraction of less than 50%, or clinically significant aortic regurgitation or mitral valve disease or if they had undergone cardiac surgery. | SAVR vs Med Rx | Composite of operative mortality or death from cardiova-scular causes | ROB2: Low |
| Leon et a. 2010; Kapadia et al. 2014 | PARTNER B | RCT | USA, Canada, Germany | 449 | TAVR: 83.0, Med Rx: 83.2 | 3 | Symptomatic, mean gradient > 40 mm Hg or jet velocity > 4.0 m/s or an aortic valve area (AVA) of < 0.8 $cm^2$ (or AVA index < 0.5 $cm^2/m^2$). Congenital unicuspid or congenital bicuspid valve was excluded. | TAVR vs Med Rx | All-cause mortality at 1 year, 2 years and 3 years | ROB2: Some concerns |
| Madhav-an et al. 2023 | SAPIEN 3 | Propensity-score matched cohort | USA, Canada | 1,566 | 81.6 | 5 | Severe, symptomatic AS AVA ≤0.8 cm2 (index 0.5 $cm^2/m^2$) or peak velocity >4 m/s or mean PG >40 mmHg. Patients were deemed to | TAVR (PARTNER 2 S3 intermedia- | Composite endpoint of death or | NOS: 8 |

| | | | | | | | | | |
|---|---|---|---|---|---|---|---|---|---|
| | | | | | | | be at intermediate risk for operative complications or death. | te-risk (P2S3i) single-arm studies) vs SAVR (PARTNER 2) | disabling stroke | |
| Kvasler-ud et al. 2021 | Norway | Retrospective cohort | Norway | 2341 | N/A | 7 | Age > 18 years, severe AS. Severe aortic stenosis was defined as an aortic valve area ≤ $1cm^2$, mean pressure gradient ≥ 40 mmHg and maximal jet velocity ≥ 4m/s. | AVR vs Med Rx | 7-year survival | NOS: 7 |
| Takeji et al. 2019 | KC registry | Retrospective propensity-score matched cohort, registry | Japan | 556 | TAVR: 84.6, Med Rx: 85.1 | 2 | Severe AS was defined as peak aortic jet velocity (Vmax) >4.0 m/s, mean aortic pressure gradient (PG) >40 mmHg, or aortic valve area (AVA) <1.0 $cm^2$. Patients on hemodialysis and those asymptomatic patients with Vmax <5m/s and LVEF ≥ 50% were excluded. | TAVR vs Med Rx | All-cause mortality | NOS: 7 |
| Taniguc-hi et al. 2015 | CURRENT registry | Retrospective propensity-score matched cohort, registry | Japan | 582 | AVR: 71.6, Med Rx: 73.1 | 5 | Severe AS (peak aortic jet velocity [Vmax] >4.0 m/s, mean aortic pressure gradient [PG] >40 mm Hg, or aortic valve area [AVA] <1.0 $cm^2$). Patients with a history of aortic valve repair/replacement/plasty or percutaneous aortic balloon valvuloplasty were excluded. | AVR vs Med Rx | All-cause mortality | NOS: 9 |
| Tourne-au et al. 2010 | Letour_STS | Retrospective cohort | USA | 694 | 71 | 10 | Patients aged 40 years or more diagnosed with asymptomatic severe AS defined by a peak systolic velocity of 4 m/s or greater. Patients with multivalvular involvement, moderate to severe aortic regurgitation, history of clinical coronary artery disease, and prior aortic valve intervention or prior cardiac surgery of any cause were excluded. | SAVR vs Med Rx | All-cause mortality | NOS: 7 |
| Clark et al. 2012 | US Medicare | Retrospective Cohort | USA | 2150 | 82 | 5 | Patients who would be candidates for TAVR based on the presence of severe, symptomatic AS who were considered to be at high risk for surgical AVR and were undergoing medical management. | Med Rx | 5-year survival | N/A |
| Génére-ux et al. 2023 | Egnite | Retrospective cohort | USA | 12129 | 78.4 | 4 | Patients with severe AS by echocardiographic reports and > 18 years of age. | Med Rx | All-cause mortality | N/A |
| Kitai et al. 2011 | Kitai | Retrospective cohort | Japan | 164 | 70 | 6 | Patients with severe AS (a maximal jet velocity ≥ 4.0 m/s, or mean aortic pressure gradient (MPG) ≥ 40 mm Hg, or an aortic valve area (AVA) <1.0 $cm^2$) and patients with very severe AS (a maximal jet velocity ≥ 5.0 m/s, or MPG ≥ 50 mm Hg or an AVA <0.6 $cm^2$) | Med Rx | 6-year survival | N/A |
| Oh et al. 2019 | Korean | Retrospective cohort | Korean | 180 | 78 | 5 | Symptomatic AS who refused undertaking AVR and treated conservatively. Patients | Med Rx | All-cause mortality | N/A |

| | | | | | | | | | | |
|---|---|---|---|---|---|---|---|---|---|---|
| | | | | | | | without symptoms, patients who underwent surgical or percutaneous AVR within 6 months interval from the diagnosis, patients with concomitant moderate to severe valvular diseases other than AS and patients with other obvious causes of developing symptoms other than AS were excluded. | | | |
| Strange et al. 2019 | NEDA | Retrospective cohort, registry | Australia | 6383 | N/A | 15 | Severe AS, characterized as either high-gradient (mean gradient >40.0 mm Hg and/or peak velocity >4.0 m/s with or without an AV area <1 cm2) or low-gradient (AV area <1 $cm^2$ in the absence of high-gradient AS) | Med Rx | All-cause mortality | N/A |
| Suzuki et al. 2018 | Suzuki | Retrospective cohort | Japan | 63 | 87 | 4 | Asymptomatic adults aged 80 and older with preserved left ventricular ejection fraction (LVEF; > 50%) with severe AS ((AVA < 1.0 $cm^2$). | Med Rx | All-cause mortality | N/A |
| Varadar-ajan et al. 2006 | California | Retrospective cohort | USA | 453 | 75 | 10 | Severe aortic stenosis defined as a valve area of 0.8 $cm^2$ or less. | Med Rx | 10-year survival | N/A |

ROB2: a revised tool for assessing risk of bias in randomized trials; NOS: The Newcastle-Ottawa Scale; AVR: aortic valve replacement; AS: aortic stenosis; Med Rx: medical therapy; PG: aortic pressure gradient; SAVR: surgical aortic valve replacement; TAVR: transcatheter aortic valve replacement.
